# The Optimal Prediction Model for Successful External Cephalic Version

**DOI:** 10.1101/2024.07.03.24309734

**Authors:** Rahul Sai Yerrabelli, Peggy K. Palsgaard, Priya Shankarappa, Valerie Jennings

## Abstract

**Objective:** The majority of breech fetuses are delivered by Cesarean birth as few physicians are trained in vaginal breech birth. An external cephalic version (ECV) can prevent Cesarean delivery and the associated morbidity in these patients. Current guidelines recommend all patients with breech presentation be offered an ECV attempt. Not all attempts are successful, and an attempt does carry some risks so shared decision-making is necessary. To aid in patient counseling, over a dozen prediction models to predict ECV success have been proposed in the last few years. However, very few models have been externally validated, and thus none have been adopted into clinical practice. This study aims to use data from a United States hospital to provide further data on ECV prediction models.

**Study Design:** This study retrospectively gathered data from Carle Foundation Hospital and used it to test six models previously proposed to predict ECV success. These models were Dahl 2021, Bilgory 2023, López Pérez 2020, Kok 2011, Burgos 2010, and Tasnim 2012 (GNK-PIMS score).

**Results:** 125 patients undergoing 132 ECV attempts were included. 69 attempts were successful (52.2%). Dahl 2021 had the greatest predictive value (AUC 0.779), while Tasnim 2012 performed the worst (AUC 0.626). The remaining models had similar predictive values as each other (AUC 0.68-0.71). Bootstrapping confirmed that all models except Tasnim 2012 had confidence intervals not including 0.5. The bootstrapped 95% AUC confidence interval for Dahl 2021 was 0.71-0.84. In terms of calibration, Dahl 2021 was well calibrated with predicted probabilities matching observed probabilities. Bilgory 2023 and López Pérez were poorly calibrated.

**Conclusion:** Multiple prediction tools have now been externally validated for ECV success. Dahl 2021 is the most promising prediction tool.

**Key Points:**

- Prediction models can be powerful tools for patient counseling
- The odds of ECV success can estimated based on patient factors and clinical findings
- Of the 6 tested models, only Dahl 2021 appears to have good predictive value and calibration

## Introduction

Breech presentation complicates approximately 3-4% of term pregnancies, and transverse lie complicates an additional 0.1%-0.2%^1–3^. The three main options for malpresentation presentation include vaginal breech birth (VBB), Cesarean delivery, or external cephalic version (ECV) followed by cephalic vaginal delivery ^1^. Vaginal breech births are extremely rare in the United States and only offered in a limited number of centers. External cephalic version is a physical maneuver where physicians press on the mother’s abdomen in the third trimester to try to rotate to a cephalic (“head down”) position. If successful, the ECV allows for a cephalic vaginal delivery and foregoes the need for a Cesarean delivery or VBB. As such, ECVs are cost effective and noninvasive procedures that decrease Cesarean rates and avoid the associated morbidity for patients^2^. Current American and British guidelines recommend that patients who are near term with breech presentation be offered an ECV if there are no contraindications^2,4,5^.

Wide variation in ECV success rates have been reported, although most estimates are usually around 50-60%^2,5–7^. Furthermore, not every patient has the same likelihood of procedure success. Some risk factors for ECV failure include low amniotic fluid volume, nulliparity, obesity, descent of the breech into the pelvis, and later gestational age^8–11^. Regardless of whether the procedure is successful, the procedure can rarely result in complications, the most severe include placental abruption (0.18%), umbilical cord prolapse (0.18%), fetomaternal hemorrhage (0.9%), rupture of membranes (0.22%), and fetal death (0.19%) ^7^. As such, it is recommended that an ECV only be performed in an institution where an emergency Cesarean delivery can be performed and that shared decision making be employed to determine whether to proceed with an ECV vs a scheduled Cesarean birth.. Given the overall risks of the procedure, there has been a significant move in the field to publish models which predict an individual patient’s success, to aide patients and practitioners in deciding for or against this intervention.

Many statistical models have been published in the last decade which predict the success of an ECV depending on a multitude of variables such as: parity, placental location, amniotic fluid volume, fetal sex, and descent of the presenting part^12^. We recently published a systematic review of the 25 models that have been published so far, 14 of which in the last 5 years. The vast majority of models had no external validation and thus were not being used in clinical practice. Because of various statistical biases, it is common for a prediction model to be overfit and appear to perform well in the original study, but be poorly predictive in reality. Thus, it is critical that rigorous external validation be performed before these models are used clinically as acting on incorrect predictions is worse than have no prediction at all. Therefore, in our systematic review, we determined that rather than new models to be developed, there is a need for validation of the existing models to determine their suitability for clinical use^13^. This study aims to fill that gap in the current literature and uses data from a hospital in the United States to externally validate six different existing prediction models for ECV success.

## Methods

### Study design

We performed a retrospective cohort study at Carle Foundation Hospital in Urbana, Illinois, USA. Approval from the institutional review board (IRB) was obtained prior to data collection and the study was deemed exempt. Inclusion criteria were pregnant individuals in whom one or more ECVs were attempted or performed. ECVs in patients with a prior Cesarean section (TOLAC) and in non-breech (e.g., transverse and oblique lie) presentations were included. Twin pregnancies were excluded. Patients were excluded if the ECV or the delivery did not happen at our institution or if the patient did not have any formal ultrasounds available in our EMR.

Data was retrieved from maternal medical records including demographic characteristics, pregnancy information, ultrasound information, the success of the ECV attempt, and delivery information. All charts were manually reviewed by the authors to confirm ECV success and final method of delivery. ECV success was defined as cephalic presentation when the physician left the room after the procedure. If the fetus returned to non-cephalic presentation after, then a second attempt was considered a separate ECV altogether. Fetal presentation was obtained from the patient’s most recent formal ultrasound before the ECV; if the patient’s last ultrasound indicated cephalic presentation, then presentation information was obtained from physician notes at the time of the ECV. REDCap was used to store the data until it was deidentified.

Demographic data was collected and tabulated. Pre-gravid BMI (BMI prior to pregnancy or at first prenatal visit) was able to be collected for almost all patients, but BMI before ECV and BMI at delivery was only available for a subset of patients. Many of the models included BMI, but only some specified which BMI was used. The appropriate BMI was used when possible; otherwise, the last available BMI was used. Some patients had other missing data (most commonly for fetal presentation); thus, these patients were only used for evaluating prediction models that did not require that variable.

All models except Dahl 2021^14^ used amniotic fluid volume; four models used amniotic fluid index (AFI) while Burgos 2010^15,16^ used qualitative terms (e.g. “scarce”, “normal”, or “abundant”). AFI was collected from patient records. For calculation of the Burgos 2010 model, qualitative criteria were converted to oligohydramnios (AFI<5), normal, and polyhydramnios (AFI≥24) ^17,18^. Five patients did not have an AFI reported, but did have a single deepest pocket (SDP); this was approximated to an AFI value by multiplying by 3.

Some models required estimated fetal weight (EFW). The last growth ultrasound was used to obtain the EFW percentile. Then, using the World Health Organization Fetal Growth Charts, the EFW at the time of ECV was calculated using the EFW percentile, gestational age at ECV, and the fetal sex^19,20^ (see https://github.com/jcarvalho45/whoFetalGrowth for code and dataset).

### Statistical analysis

Six prediction models were chosen from the prior systematic review: Dahl 2021^14^, Bilgory 2023^21,22^, López Pérez 2020^12^, Kok 2011^23^, Burgos 2010^15,16^, and Tasnim 2012 (GNK-PIMS score)^24^. These models were chosen because they did not require any additional measurements (e.g., forebag size, cervical exam before ECV) beyond what is routinely already performed at our institution. The score or predicted probability was calculated for each model. The receiver operating characteristic (ROC) curves were calculated for each model by determining the sensitivity and specificity values at each threshold. The area under the curve (AUC) was calculated and used as a predictor of discriminative power, and this was compared between the models.

Because the ROC does not naturally have a method to determine confidence intervals, bootstrapping statistical techniques were used to simulate 1000 new datasets from our original dataset by sampling with replacement. This was done for each model and the simulations were used to create confidence intervals for the AUC as well as the ROC curves themselves.

Finally, calibration plots were calculated to give a measure of how well the calculated probabilities compared to the observed outcomes^25^. This was performed by calculating the odds of success at each decile of predicted probability (e.g., in cases with a predicted probability of 70-79%, what percent of the time was the ECV actually successful?). Quality of calibration was assessed visually.

Calculations were performed with Python version 3.10 using numpy, pandas, and sklearn (scikit-learn) libraries and in Jupyter notebooks hosted by Google colab. Figures were created using plotly 5.15. The code is hosted on GitHub and will be published online after publication of this study. The underlying data was deidentified and cleaned for publication with this manuscript for reproduction or for future analyses.

## Results

### Population demographics

We ultimately obtained data from 125 patients who underwent a total of 132 ECV attempts, of which 69 were successful (52.2%) as seen in **Table 1** and **Table 2**. 53 of the patients ultimately had a vaginal delivery including 50 of those with successful ECVs and 3 of those with failed ECVs who spontaneously verted afterwards. No patients had vaginal breech births. 107 patients were breech, 15 patients were transverse, and 3 were oblique before the first ECV attempt. Our sample had a largely white population, which is consistent with the demographics of our geographical region. No patients had oligohydramnios and six patients had polyhydramnios (AFI ≥24), of which five had mild polyhydramnios (AFI<30) and five had successful ECVs (all except the patient with moderate polyhydramnios). 1 patient did not have any recorded BMI values. 116 patients did not have a recorded EFW percentile. Only 40 patients had information of breech type (e.g. frank, complete, incomplete). The underlying data from this study is given in **Supplement 1**.

**Table 1.** Demographic information and other baseline characteristics.

|  | Count | % |
| --- | --- | --- |
| ***ECVs Performed*** |  |  |
| 1 | 121 | 97% |
| 2 | 2 | 2% |
| 3 | 1 | 1% |
| 4 | 1 | 1% |
| ***Parity*** |  |  |
| 0 | 54 | 43% |
| 1 | 32 | 26% |
| 2 | 24 | 19% |
| 3 | 10 | 8% |
| ≥4 | 5 | 4% |
| ***Race*** |  |  |
| White | 85 | 69% |
| Black or African American | 21 | 16% |
| Asian | 11 | 9% |
| American Indian / Alaskan Native | 1 | 1% |
| Patient refused | 1 | 1% |
| All other/none of the above | 14 | 11% |
| ***Ethnicity*** |  |  |
| Not Hispanic or Latino | 111 | 89% |
| Hispanic or Latino | 13 | 10% |
| Unknown/patient refused | 1 | 1% |
| ***Fetal Sex*** |  |  |
| Female | 68 | 54% |
| Male | 57 | 46% |
| ***Delivery*** |  |  |
| **Cesarean - Total** | 82 | 64% |
| Low transverse, primary | 69 | 53% |
| Low transverse, repeat | 7 | 5% |
| Low transverse, unspecified | 5 | 4% |
| Vertical, primary | 1 | 1% |
| **Vaginal - Total** | 47 | 36% |
| Vacuum-assisted | 3 | 2.4% |
| Spontaneous | 44 | 34% |
| ***Presentation / Lie*** |  |  |
| **Breech - Total** | **107** | **86%** |
| Frank | 25 | 20% |
| Footling | 7 | 6% |
| Complete | 7 | 6% |
| Incomplete | 1 | 1% |
| Other/unknown breech | 67 | 54% |
| **Transverse** | **15** | **12%** |
| **Oblique** | **3** | **2%** |

**Table 2.** Baseline characteristics continued.

| Variable | Mean ± SD | Min - Max |
| --- | --- | --- |
| Maternal age (yr) | 30.2 ± 4.9 | 18 - 40 |
| Initial BMI (kg/m<sup>2</sup>) | 28.2 ± 6.7 | 17.6 - 46.3 |
| Last BMI (kg/m<sup>2</sup>) | 31.1 ± 6.3 | 19.9 - 47.3 |
| GA at ECV (wk) | 37.9 ± 0.92 | 36w 4d - 40w 6d |
| GA at delivery (wk) | 39.3 ± 0.86 | 37w 1d - 41w 3d |
| ECV to delivery time (days) | 9.6 ± 7.34 | 0 - 27 |
| AFI | 14.7 ± 4.96 | 5.73 - 34.97 |
| EFW at last US (percentile), n=112 | 46.8 ± 19.9 | 9 - 95 |
| EFW, calculated at ECV (g), n=112 | 3121 ± 298 | 2597 - 4277 |

### Available models – discrimination ability

The following 6 models were studied: Dahl 2021^14^, Bilgory 2023^21,22^, López Pérez 2020^12^, Kok 2011^23^, Burgos 2010^15,16^, and Tasnim 2012 (GNK-PIMS score)^24^. **Table 3** outlines the varying characteristics of these models including the AUC. In our population, all models had at least some predictive value (AUC >0.5), with Dahl 2021 having the greatest AUC at 0.78 (**Figure 1**). Bilgory 2023^21,22^, López Pérez 2020^12^, Kok 2011^23^, and Burgos 2010^15,16^ all had similar predictive values with AUC 0.68-0.71. Tasnim 2012^24^ had the lowest predictive value in our study with an AUC of 0.63. When bootstrapping was performed to simulate confidence intervals for the AUC, Tasnim 2012^24^ was not statistically significant with 0.5 being inside the confidence interval (**Figure 2**). However, the confidence interval was wide for Tasnim 2012 because the majority of our data did not have fetal presentation (e.g., frank vs complete vs incomplete breech) recorded and thus was not included in the analysis of Tasnim 2012. The confidence intervals for the AUC overlapped between the majority of the models so confirmation of which model had the best discriminative ability was not able to be done. A bootstrapped ROC curve is shown for Dahl 2021 in **Figure 3** and a table of bootstrapped AUC value for all the models is shown in **Supplement 2**.

**Figure 1.**
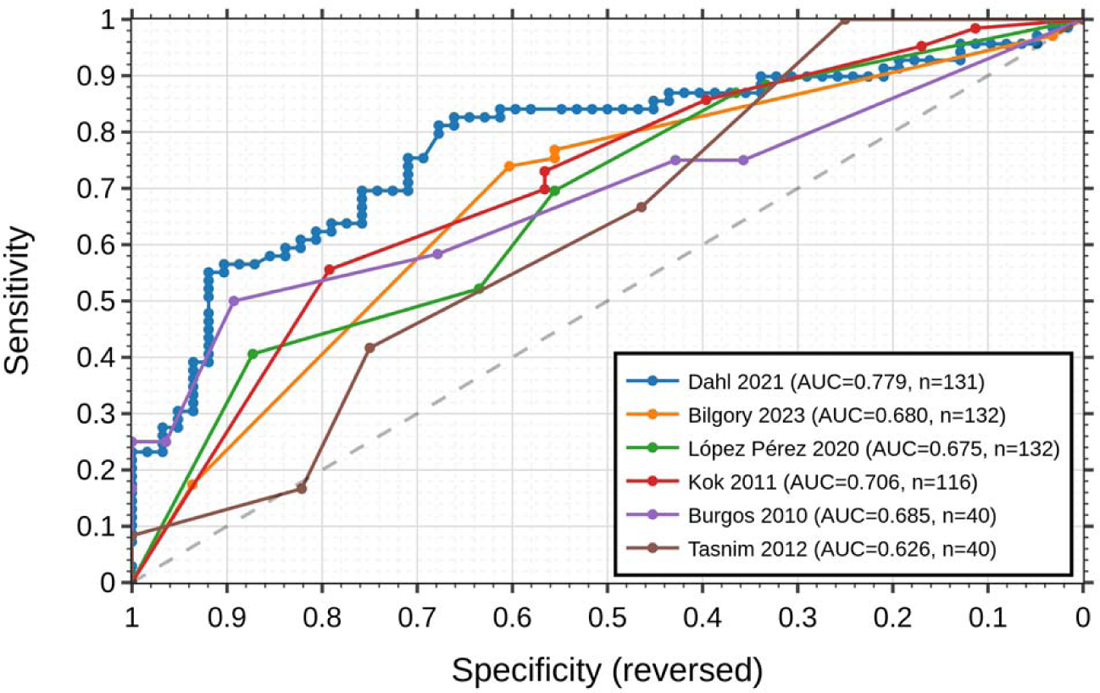
ROC curves of all models. The predictive ability of all the models is displayed using receiver operating characteristic (ROC) curves and the area under the curve (AUC).

**Figure 2.**
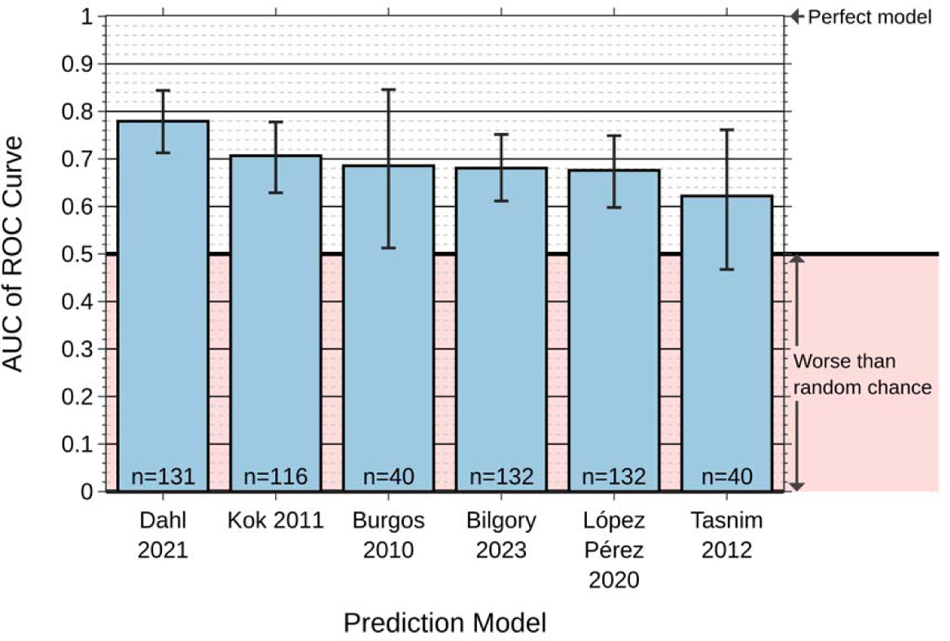
AUC of models with bootstrapping. The AUC of all the models is displayed as a bar chart for comparison. 95% confidence intervals were estimated using bootstrapping from 1,000 simulated datasets

**Figure 3.**
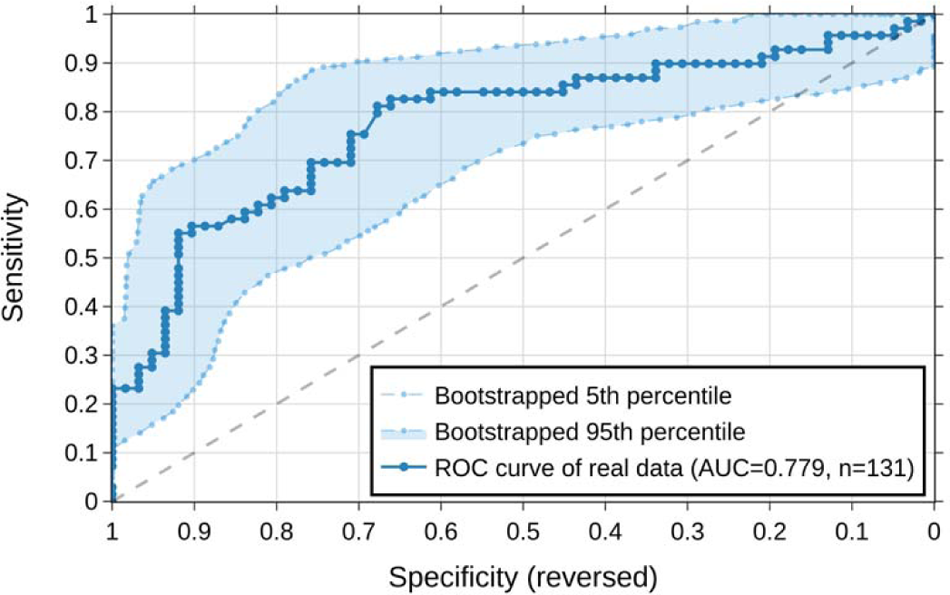
ROC curve for Dahl 2021 with bootstrapping

**Table 3.**
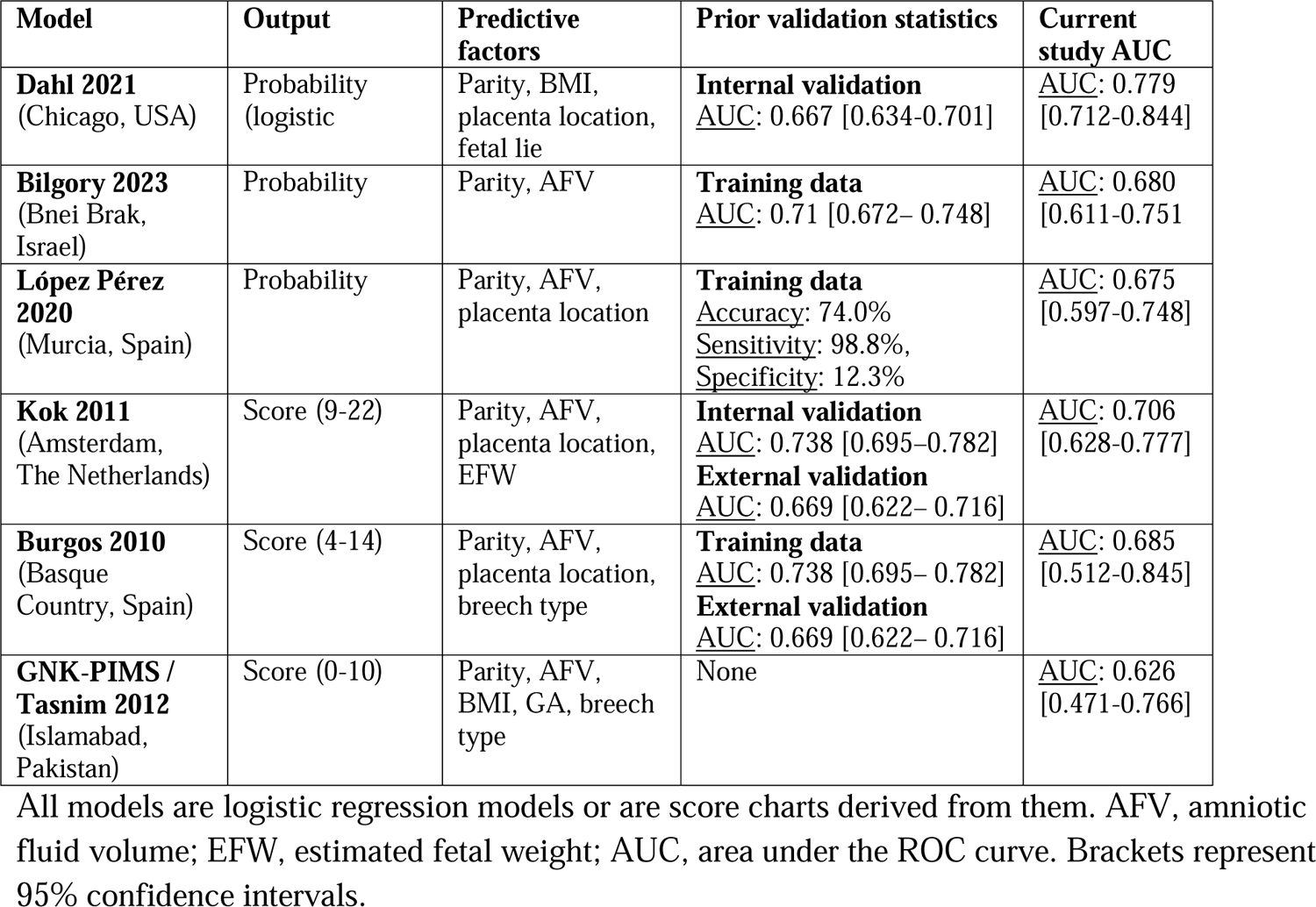
Background on prediction models.

### Available models – calibration

Dahl 2021^14^ was relatively well calibrated in our population (**Figure 4**). Bilgory 2023^21, 2212^ had poor calibration without a specific tendency to over or under calibrate. López Pérez 2020 was consistently under-calibrated. That is, it would consistently underestimate the probability of success in our population by a relatively fixed amount. The remaining three models return score values instead of predicted probabilities. Thus, calibration curves cannot be properly assessed. A probability of success by score value is shown for Kok 2011, Burgos 2010, and Tasnim 2012 in **Figure 5**. The probability of success poorly correlated with score for Kok 2011 and Tasnim 2012, but correlated well for Burgos 2010.

**Figure 4.**
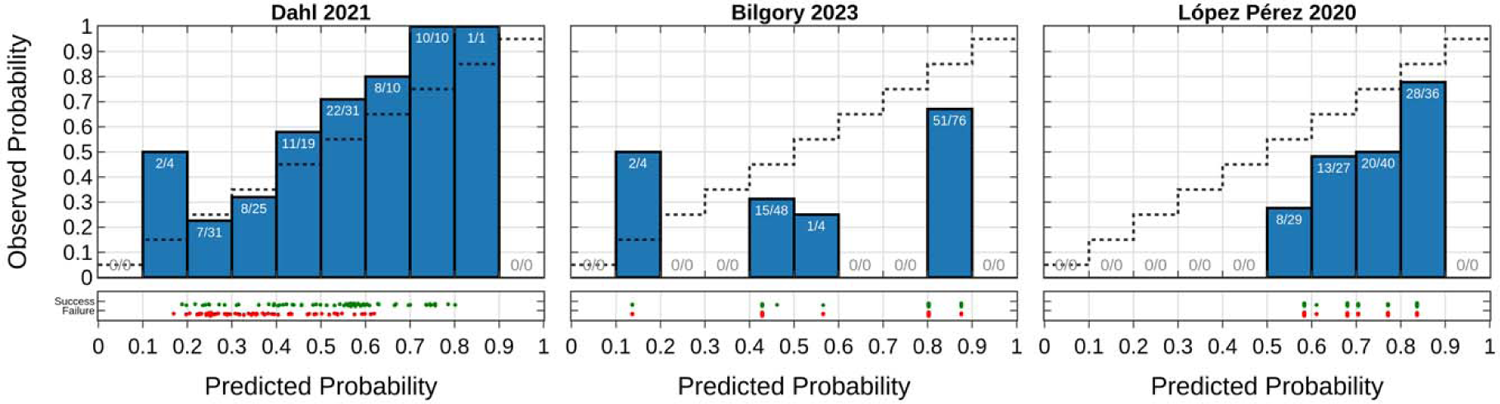
Calibration curves. The calibration is displayed for the models that output probabilities: Dahl 2021, Bilgory 2023, and López Pérez 2020. This figure shows how well the predicted probabilities match the observed probability. Only Dahl 2021 has a good match.

**Figure 5.**
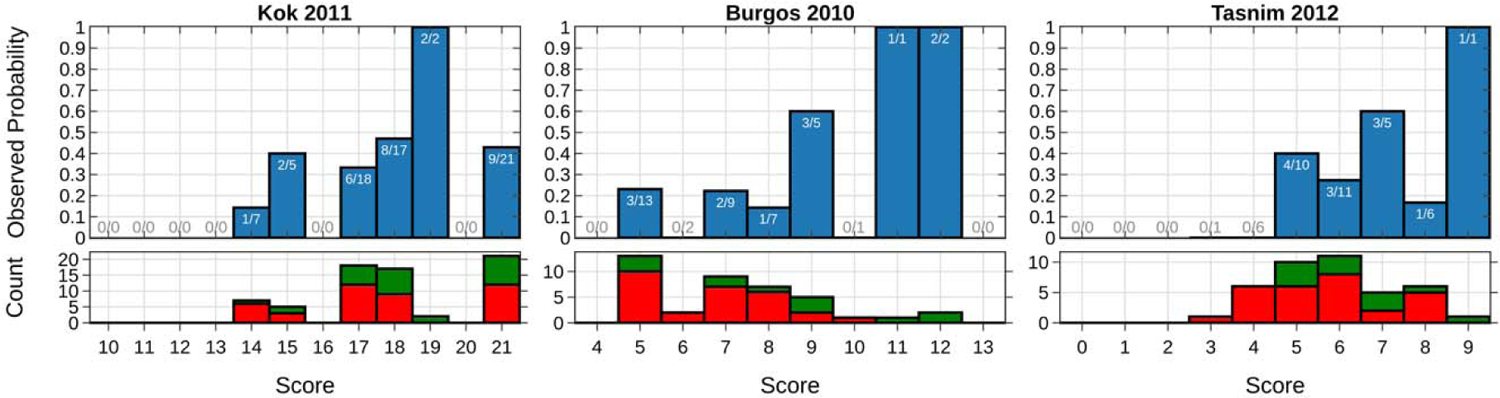
Observed probability by predicted score. This is shown as an alternative to the calibration curve for the models (Kok 2011, Burgos 2010, Tasnim 2012) that do not output a probability score.

## Discussion

The prediction of ECV success has the potential to be an incredibly powerful tool in counseling patients as they weigh the risks and benefits of the procedure. Prediction models have the opportunity to give specific, personalized information to patients. However, they have the ability to cause harm if they are inaccurate or poorly calibrated. In reality, confidently providing a patient an inaccurate odds of success is worse than providing no prediction at all.

Unfortunately, there has not been a clinical consensus on the optimal tool despite dozens of models having been proposed. Our prior systematic review identified lack of external validation studies as a major gap in the literature, prohibiting clinical use of published models.

In this article, we set out to validate the 6 models that could be used in our population as they used variables which are already collected as part of routine care at our institution. These models are Dahl 2021^14^, Bilgory 2023^21,22^, López Pérez 2020^12^, Kok 2011^23^, Burgos 2010^15,16^, and Tasnim 2012 (GNK-PIMS score)^24^. Our data demonstrates that all of these models have at least some predictive value, with the exception of Tasnim 2012 (which was not statistically significant by bootstrapping, likely because of the small number of patients with sufficient recorded data to calculate this model). Of all the models, Dahl 2021 had the greatest AUC, although the bootstrapped confidence interval still overlapped with the other models. Of the three models that output a specific probability, only Dahl 2021 appeared overall well calibrated. Based off the available evidence from this study, Dahl 2021 appears to overall be the best model for clinical use.

This is the first external validation study of Dahl 2021. While it is generally expected that external validation studies will show worse predictive values than the original study because of some degree of unavoidable statistical bias, the opposite was true here. Dahl 2021’s AUC in our study was 0.779 (confidence interval of 0.712-0.844), which is higher than the original study, which showed an AUC of 0.667 (confidence interval of 0.634-0.701). Overall, the results are very promising for Dahl 2021.

There are some limitations with our study. Due to the retrospective nature of our study, we only evaluated models that used variables routinely measured in our institution. Our institution does not routinely measure uterine tone^26,27^, cervical exam/fetal station^28–30^ and forebag size^31^, which are components of other models. One of these models may in fact have greater predictive value than the ones we evaluated in our study. On the other hand, these models would also be less practical to implement in clinical practice because they would require measuring additional variables. Additionally, these variables would not be known until close to ECV; thus, it would be hard to counsel patients in advance using one of these models. Since many models required cervical exam or fetal station, we suggest future research study be a prospective trial where cervical exam is performed.

## Declarations

### Ethics approval and consent to participate

No new unpublished data from humans or other animals was included in this study.

### Consent for publication

Not applicable.

## Data Availability

All data produced in the present work are contained in the manuscript and supplement.

## Acknowledgments

We thank artificial intelligence engineer Benjamin M. Pikus for his help in deciding how to evaluate prediction model performance.

## Supplement

**Supplement 1 Deidentified patient data to reproduce the analyses**. For yes/no questions, 0: no, 1: yes. For baby gender, 1: female, 2: male. For ethnicity, 11: Hispanic, 12: not Hispanic, 13: unknown/patient refused. For language, 1: English, 2: French, 3: Spanish, 15: Portuguese. For patient race, 1: White, 2: Black, 3: none of the above/all other races, 4: Hispanic, 5: Asian, 6: American Indian or Alaskan Native, 7: Pacific Islander or Native Hawaiian, 8: Patient refused. EFW at ECV and at delivery were calculated using the percentile from the most recent ultrasound as described in the methods. ECV success for the first, second, third, and fourth ECV attempts is given by the variables ecv_successful_1, ecv_successful_2, etc, respectively. Maternal age at delivery (age_delivery) is given in years. Gestational ages are given in days. EFWs are given in grams. Patient weights and total weight gain (twg) is given in pounds, while BMI is given in kg/m2

**Supplement 2.** Area under the curve data with bootstrapping. AUC values >0.5 are highlighted

|  | Original Data |  | 1000 Bootstrapped Simulations |  |  |  |  |  |  |  |  |
| --- | --- | --- | --- | --- | --- | --- | --- | --- | --- | --- | --- |
|  |  |  | Center |  | Percentiles |  |  |  |  |  |  |
|  | n | AUC | AUC Mean | AUC SD | AUC Min | AUC 1% | AUC 5% | AUC Median | AUC 95% | AUC 99% | AUC Max |
| Dahl 2021 | 131 | 0.7791 | 0.7794 | 0.0414 | 0.6496 | 0.6797 | 0.7124 | 0.7817 | 0.8437 | 0.8692 | 0.8891 |
| Bilgory 2023 | 132 | 0.6800 | 0.6805 | 0.0433 | 0.5581 | 0.5764 | 0.6108 | 0.6802 | 0.7510 | 0.7723 | 0.8310 |
| López Pérez 2020 | 132 | 0.6754 | 0.6760 | 0.0464 | 0.5161 | 0.5644 | 0.5972 | 0.6775 | 0.7482 | 0.7722 | 0.7960 |
| Kok 2011 | 116 | 0.7063 | 0.7066 | 0.0456 | 0.5693 | 0.5978 | 0.6282 | 0.7089 | 0.7773 | 0.8018 | 0.8389 |
| Burgos 2010 | 40 | 0.6845 | 0.6854 | 0.1021 | 0.2396 | 0.4273 | 0.5116 | 0.6859 | 0.8449 | 0.8899 | 0.9436 |
| Tasnim 2012 | 40 | 0.6265 | 0.6221 | 0.0879 | 0.3177 | 0.4121 | 0.4718 | 0.6225 | 0.7656 | 0.8219 | 0.8860 |

## References

1. Richmond AK, Ashworth JR. Management of malposition and malpresentation in labour. Obstet Gynaecol Reprod Med. 2023;33(11):325–333. doi:10.1016/j.ogrm.2023.08.004

2. External Cephalic Version: ACOG Practice Bulletin, Number 221. Obstet Gynecol. 2020;135(5):e203–e212. doi:10.1097/AOG.0000000000003837

3. Hill LM. Prevalence of breech presentation by gestational age. Am J Perinatol. 1990;7(1):92–93. doi:10.1055/s-2007-999455

4. ACOG Committee Opinion No. 745: Mode of Term Singleton Breech Delivery. Obstet Gynecol. 2018;132(2):e60–e63. doi:10.1097/AOG.0000000000002755

5. External Cephalic Version and Reducing the Incidence of Term Breech Presentation. BJOG Int J Obstet Gynaecol. 2017;124(7):e178–e192. doi:10.1111/1471-0528.14466

6. Nalam RL, Chinnachamy P, Emmanuel P. External Cephalic Version: A Dying Art Worth Reviving. J Obstet Gynecol India. 2018;68(6):493–497. doi:10.1007/s13224-018-1090-z

7. Grootscholten K, Kok M, Oei SG, Mol BWJ, van der Post JA. External cephalic version-related risks: a meta-analysis. Obstet Gynecol. 2008;112(5):1143–1151. doi:10.1097/AOG.0b013e31818b4ade

8. Ben-Meir A, Erez Y, Sela HY, Shveiky D, Tsafrir A, Ezra Y. Prognostic parameters for successful external cephalic version. J Matern-Fetal Neonatal Med Off J Eur Assoc Perinat Med Fed Asia Ocean Perinat Soc Int Soc Perinat Obstet. 2008;21(9):660–662. doi:10.1080/14767050802244938

9. Ebner F, Friedl TWP, Leinert E, et al. Predictors for a successful external cephalic version: a single centre experience. Arch Gynecol Obstet. 2016;293(4):749–755. doi:10.1007/s00404-015-3902-z

10. Kok M, Cnossen J, Gravendeel L, van der Post J, Opmeer B, Mol BW. Clinical factors to predict the outcome of external cephalic version: a metaanalysis. Am J Obstet Gynecol. 2008;199(6):630.e1-7; discussion e1-5. doi:10.1016/j.ajog.2008.03.008

11. Chaudhary S, Contag S, Yao R. The impact of maternal body mass index on external cephalic version success. J Matern-Fetal Neonatal Med Off J Eur Assoc Perinat Med Fed Asia Ocean Perinat Soc Int Soc Perinat Obstet. 2019;32(13):2159–2165. doi:10.1080/14767058.2018.1427721

12. López-Pérez R, Lorente-Fernández M, Velasco-Martínez M, Martínez-Cendán JP. Prediction model of success for external cephalic version. Complications and perinatal outcomes after a successful version. J Obstet Gynaecol Res. 2020;46(10):2002–2009. doi:10.1111/jog.14385

13. Yerrabelli RS, Lee C, Palsgaard PK, Lauinger AR, Abdelsalam O, Jennings V. Prediction Models for Successful External Cephalic Version: An Updated Systematic Review. Am J Perinatol. Published online January 12, 2024. doi:10.1055/a-2211-4806

14. Dahl CM, Zhang Y, Ong JX, et al. A Multivariable Predictive Model for Success of External Cephalic Version. Obstet Gynecol. 2021;138(3):426–433. doi:10.1097/AOG.0000000000004518

15. Burgos J, Melchor JC, Pijoán JI, Cobos P, Fernández-Llebrez L, Martínez-Astorquiza T. A prospective study of the factors associated with the success rate of external cephalic version for breech presentation at term. Int J Gynecol Obstet. 2011;112(1):48–51. doi:10.1016/j.ijgo.2010.07.023

16. Burgos J, Cobos P, Rodriguez L, et al. Clinical score for the outcome of external cephalic version: A two-phase prospective study: Clinical score for external cephalic version. Aust N Z J Obstet Gynaecol. 2012;52(1):59–61. doi:10.1111/j.1479-828X.2011.01386.x

17. Reddy UM, Abuhamad AZ, Levine D, Saade GR. Fetal Imaging: Executive Summary of a Joint Eunice Kennedy Shriver National Institute of Child Health and Human Development, Society for Maternal-Fetal Medicine, American Institute of Ultrasound in Medicine, American College of Obstetricians and Gynecologists, American College of Radiology, Society for Pediatric Radiology, and Society of Radiologists in Ultrasound Fetal Imaging Workshop. Obstet Gynecol. 2014;123(5):1070–1082. doi:10.1097/AOG.0000000000000245

18. Society for Maternal-Fetal Medicine (SMFM). Electronic address, Dashe JS, Pressman EK, Hibbard JU. SMFM Consult Series #46: Evaluation and management of polyhydramnios. Am J Obstet Gynecol. 2018;219(4):B2–B8. doi:10.1016/j.ajog.2018.07.016

19. Kiserud T, Piaggio G, Carroli G, et al. The World Health Organization Fetal Growth Charts: A Multinational Longitudinal Study of Ultrasound Biometric Measurements and Estimated Fetal Weight. PLoS Med. 2017;14(1). doi:10.1371/journal.pmed.1002220

20. Kiserud T, Benachi A, Hecher K, et al. The World Health Organization fetal growth charts: concept, findings, interpretation, and application. Am J Obstet Gynecol. 2018;218(2):S619–S629. doi:10.1016/j.ajog.2017.12.010

21. Bilgory A, Minich O, Shvaikovsky M, Gurevich G, Lessing JB, Olteanu I. Predictive Factors for Successful Vaginal Delivery after a Trial of External Cephalic Version: A Retrospective Cohort Study of 946 Women. Am J Perinatol. 2023;40(15):1679–1686. doi:10.1055/s-0041-1739505

22. Bilgory A, Minich O, Shvaikovsky M, Gurevich G, Lessing JB, Olteanu I. Erratum: Predictive Factors for Successful Vaginal Delivery after a Trial of External Cephalic Version: A Retrospective Cohort Study of 946 Women. Am J Perinatol. Published online January 4, 2022. doi:10.1055/s-0041-1742110

23. De Hundt M, Vlemmix F, Kok M, et al. External Validation of a Prediction Model for Successful External Cephalic Version. Am J Perinatol. 2012;29(03):231–236. doi:10.1055/s-0031-1285098

24. Tasnim N, Mahmud G, Javaid K. GNK-PIMS Score: A Predictive Model for Success of External Cephalic Version. J South Asian Fed Obstet Gynaecol. 2012;4(2):99–102. doi:10.5005/jp-journals-10006-1184

25. Van Calster B, Nieboer D, Vergouwe Y, De Cock B, Pencina MJ, Steyerberg EW. A calibration hierarchy for risk models was defined: from utopia to empirical data. J Clin Epidemiol. 2016;74:167–176. doi:10.1016/j.jclinepi.2015.12.005

26. Anand K, Keepanasseril A, Amala R, Nair NS. Development and validation of a clinical score to predict the probability of successful procedure in women undergoing external cephalic version. J Matern-Fetal Neonatal Med Off J Eur Assoc Perinat Med Fed Asia Ocean Perinat Soc Int Soc Perinat Obstet. 2019;34(18):2925–2931. doi:10.1080/14767058.2019.1674803

27. Wong WM, Lao TT, Liu KL. Predicting the success of external cephalic version with a scoring system. A prospective, two-phase study. J Reprod Med. 2000;45(3):201–206.

28. Zheng LG, Zhang HL, Chen RX, et al. Scoring system to predict the success rate of external cephalic versions and determine the timing of the procedure. Eur Rev Med Pharmacol Sci. 2021;25(1):45–55. doi:10.26355/eurrev_202101_24345

29. Hutton EK, Simioni JC, Thabane L. Predictors of success of external cephalic version and cephalic presentation at birth among 1253 women with non-cephalic presentation using logistic regression and classification tree analyses. Acta Obstet Gynecol Scand. 2017;96(8):1012–1020. doi:10.1111/aogs.13161

30. Newman RB, Peacock BS, Peter VanDorsten J, Hunt HH. Predicting success of external cephalic version. Am J Obstet Gynecol. 1993;169(2):245–250. doi:10.1016/0002-9378(93)90071-P

31. Isakov O, Reicher L, Lavie A, Yogev Y, Maslovitz S. Prediction of Success in External Cephalic Version for Breech Presentation at Term. Obstet Gynecol. 2019;133(5):857–866. doi:10.1097/AOG.0000000000003196

